# Static and dynamic intracerebral signal analysis reveals protective networks against seizures in drug-resistant focal epilepsy

**DOI:** 10.1101/2025.01.20.25320855

**Authors:** Roberta Di Giacomo, Pablo Núñez, Jesús Poza, Victor Rodríguez-González, Carlos Gómez, Alessandra Burini, Laura Castana, Marco de Curtis, Laura Tassi, Giulia Varotto

**Affiliations:** Epilepsy Unit, Fondazione IRCCS Istituto Neurologico Carlo Besta, Milan, Italy; Coma Science Group, GIGA-Consciousness, University of Liège, Liège, Belgium; Biomedical Engineering Group, University of Valladolid, Valladolid, Spain; Centro de Investigación Biomédica en Red de Bioingeniería, Biomateriales y Nanomedicina (CIBER-BBN), Spain; IMUVA, Instituto de Investigación en Matemáticas, University of Valladolid, Valladolid, Spain; Neurology Unit, Department of Medicine (DMED), University of Udine, Udine, Italy; Claudio Munari Epilepsy Surgery Centre, Niguarda Hospital, Piazza Ospedale Maggiore 3, 20162 Milan, Italy; Laboratory for Clinical Neuroscience, Center for Biomedical Technology, Universidad Politécnica de Madrid, Madrid, Spain

**Keywords:** drug-resistant epilepsy, stereo-electroencephalography, brain networks, functional connectivity, epileptogenic zone

## Abstract

**Background:** Epilepsy research increasingly emphasizes the role of brain network dynamics in seizure generation and propagation. This study explores static and dynamic functional networks in subjects with drug-resistant epilepsy, to identify mechanisms that enhance or inhibit seizure initiation. To this aim, we analyzed functional connectivity of brain networks preceding ictal minor electrical discharges and major seizures in epileptogenic and non-epileptogenic zones explored with intracerebral electrodes.

**Material and methods:** Stereo-electroencephalographic signals were recorded from 39 patients with focal drug-resistant epilepsy during presurgical monitoring. Static functional connectivity was analyzed using graph theory metrics, whereas dynamic connectivity through the analysis of the complexity and dwell times of brain meta-states activations.

**Results:** Static connectivity analysis revealed significant alterations in network centrality, integration, and segregation properties, with distinct patterns observed in resting conditions just ahead minor electrical discharges and major seizures. Specifically, network analysis before minor electrical discharges exhibited increased nodal strength and reduced betweenness centrality in the epileptogenic zone, associated with increased integration and reduced segregation in non-epileptogenic zones. Dynamic connectivity analysis showed lower complexity and longer stability of meta-states before minor electrical discharges, particularly in high-frequency signals of non-epileptogenic zones.

**Conclusions:** Our findings provide novel and valuable insights into the dynamic reconfiguration of brain networks before epileptic seizures, suggesting an inhibitory/protective mechanism mainly involving the non-epileptogenic zones. Understanding these network changes is pivotal for improving epilepsy treatment strategies targeting dynamic network alterations.

## 1. Introduction

The unpredictability of the appearance and recurrence of seizures leads to significant impact in the life of people with epilepsy. Understanding pre-ictal electrophysiological alterations could offer valuable information to better understand seizure-related mechanisms, and to develop proper tools for seizure prediction aimed at controlling seizure occurrence. Human brain functions are increasingly understood and explained in terms of large-scale complex brain networks that dynamically evolve across multiple spatial and temporal scales. Epilepsy and several others neurological diseases have been associated with abnormal modifications of these networks.^1,2^ How these alterations lead to seizure generation and spreading is still unknown, and this information could be clinically relevant to support the epileptogenic zone (EZ) localization and to predict seizures occurrence.

Despite the large body of studies on functional connectivity (FC) in epilepsy, the understanding of the mechanisms ruling epileptogenic networks remains elusive.^3^ Most of these studies focus on *static functional connectivity* (sFC), which assumes temporal stationarity of the statistical interdependencies between signals recorded in different brain regions, thus ignoring the temporal evolution of these networks. As neuronal oscillations evolve very quickly in time,^4^ FC also reflects these rapid changes, exhibiting a high dynamic variability in time and space.^5^ The field of *dynamic functional connectivity* (dFC)^1,6^ paved the way to assess the fast fluctuations of neural networks, allowing to take full advantage of the high temporal resolution of electrophysiological data.

In this study, we used a comprehensive approach encompassing both sFC and dFC analyses. First, we used graph theory methods to quantify the topological sFC differences between different conditions. Second, we applied a dFC-based methodology to detect brain network configurations recurrent over time (referred to as “meta-states”) that act as attractors of the neural network. ^7,8^ Recently, there has been a growing interest in the study of these recurrent brain states.^5,8–10^ This dynamic approach for assessing the temporal architecture of functional connectivity has been employed for assessing the alterations associated with different neurological and psychiatric disorders. In the field of epilepsy, functional magnetic resonance imaging (fMRI) was applied either to compare brain networks states in epileptic patients and healthy controls (Abreu et al., 2019; Klugah-Brown et al., 2019) or to assess the effect of intracerebral electrical stimulation.^11^ Despite its wide use, fMRI signals have a reduced temporal resolution compared to electrophysiological data, which limits the analysis of fast brain dynamics. Conversely, electroencephalography and magnetoencephalography (MEG) allow to capture physiological and pathophysiological activity on various time scales with a high temporal resolution, although requiring more sophisticated time-series-analysis techniques.^12^ In this regard, slower state transitions and reduced activation complexity were observed in patients with schizophrenia.^8^

The gold-standard procedure to identify the brain areas responsible for seizure generation in drug-resistant epileptic (DRE) patients is the stereo-electroencephalography (SEEG); this diagnostic method is utilized during pre-surgical evaluation to identify the cortical areas to be resected to cure the patients.^13,14^ These invasive intracerebral recordings are performed when no lesion is detected with neuroimaging techniques, or when the boundaries of the lesion cannot be clearly outlined.^14,15^ SEEG reveals interictal and ictal activities that, together with semiology and anatomical data, define the epileptogenic zone (EZ, *i.e.*, the cortical areas indispensable for generation and primary organization of the seizure discharge).^13,16,17^ Moreover, SEEG represents a unique tool to localize and identify interictal activities in the lesional zone (*i.e.*, cortical area where background activity is altered and slow waves are predominant) and the irritative zone (*i.e.*, site of spiking activity).

The EZ can generate full-blown seizures as well as minor electrical discharges (MED).^18–21^ These discharges differ from the ongoing interictal background activity; visually, MED show an electrical pattern similar to the seizure onset generated in the same cortical area during major seizures (MS).^22,23^ Estimated using SEEG microelectrodes, MED have very short duration and minimal (if any) clinical correlates ^18^. MED can spread to the same regions of MS or they can involve different areas compared to MS. The co-localization of MED and MS correlates with the postsurgical outcome in patients who underwent resective surgery.^20–23^

The evolution pattern and the differences between MED and MS have been poorly investigated. Understanding the differences in terms of static and dynamic networks that precede MED *vs.* MS could offer novel insights onto seizure-generating mechanisms, especially to comprehend how brain networks reorganization either promotes or hampers seizure emergence and propagation. According to the “pull-push theory”^24,25^ – a theory suggesting a competitive interaction between synchronizing and desynchronizing brain regions to constrain seizure spreading –, we hypothesized the existence of a protective condition able to prevent the evolution of MED into MS, based on a dynamic network rearrangement involving brain regions beyond the EZ. Starting from the evidence that the SEEG patterns of both MED and MS are similar, we considered MED as a suitable model to provide a more comprehensive explanation of the synchronization/desynchronization mechanisms responsible for fostering or suppressing MS generation. To this aim, we evaluated differences in the static and dynamic FC of intracerebral networks preceding both MED and MS onsets and compared these to resting state activity observed during the interictal condition.

## 2. Materials and Methods

### 2.1. Subjects

Patients with DRE undergoing long-term SEEG recordings during the pre-surgical work-up were retrospectively selected from the clinical database collected at the *Claudio Munari* Epilepsy Surgery Centre of the Niguarda Hospital in Milan, over a 17-year historical extension (2005–2022). Database collection for research purposes was approved by the local Ethics Committee and informed consent was signed by all patients. We selected patients with: (i) both MS and MED (ii) available recordings of a sixty-second epoch free from epileptiform discharges before these events; (iii) MS starting with a sequence of events similar to MED were included in the study and (iv) at least one-year of follow-up evaluated with Engel’s seizure outcome scale. We selected patients with seizure freedom after surgery or radiofrequency SEEG-guided thermocoagulation, or a worse outcome only if due to limited surgery for functional reasons

### 2.2. SEEG recording and preprocessing

For SEEG video-monitoring, a Nihon Kohden System with 192 channels recorded at a sampling rate of 1000 Hz was used. SEEG was tailored to the patients’ individual anatomic and electro-clinical characteristics.^14^ Multi-lead electrodes with 5–18 recording sites (2mm in length and 1.5mm apart; Dixi; Besançon, France) were intracerebrally inserted according to the standard diagnostic protocol.^14^ Wake and sleep recordings were visually examined by two neurologists (RDG and AB) to define the MED and MS onset and to delineate leads included in the EZ. MEDs were not associated to obvious clinical manifestations and were identified as low-voltage fast activity discharges, often associated with very slow potentials, that could gradually develop into a rhythmic theta-delta discharge (Fig. 1), followed by electrical depression and/or slow waves. Consensus on the identification of both MED and MS, and EZ boundaries was obtained between both professionals, solving the divergences with open discussions. Fig. 1 illustrates representative SEEG signals recorded during MED and MS from one patient. Sixty seconds preceding the onset of both MED (defined hereafter as pre-MED – pMED) and MS (pMS) during the waking state were selected for each patient. Additionally, for each subject, 60 seconds of resting-state recording was selected as baseline condition at least ten minutes away from any type of ictal discharge. Leads located within the white matter, identified through MRI and electrical activity, were excluded from the analysis. Leads with artifacts were visually identified and excluded from the analysis.

**Figure 1.**
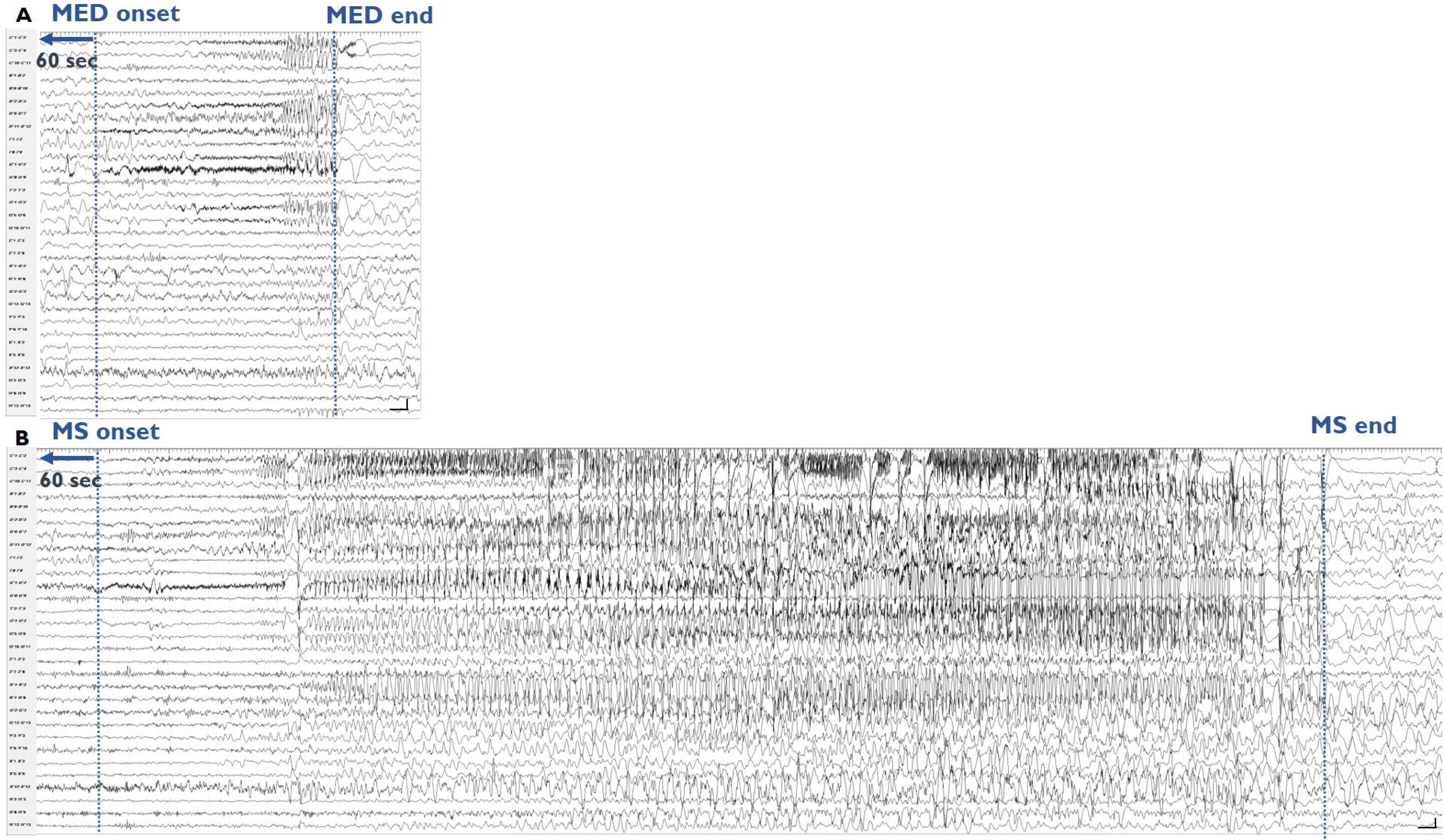
SEEG recordings (subset of selected 30 channels) of MED and MS for a representative patient. Panel A shows a MED, lasting 12 seconds involving mainly temporo-mesial, temporo-polar and insular regions. Panel B shows a MS in the same patient, lasting 62 seconds, on the same MED regions in addition to orbito-frontal leads. Calibration bars in SEEG recordings: vertical line 100 µV, horizontal line 1 s.

All recordings were down-sampled to 250 Hz to reduce computational requirements. The recordings were then filtered in the *delta* (1-4 Hz), *theta* (4-8 Hz), *alpha* (8-13 Hz), *beta1* (13-19 Hz), *beta2* (19-30 Hz), and *gamma* (30-48 Hz) bands by means of finite impulse response (FIR) filters, with both forward and backward filtering.

### 2.3. Static Functional Connectivity (sFC)

The sFC was estimated by means of the amplitude envelope correlation (AEC),^26^ due to its robustness and high reproducibility.^27^ For each of the three conditions (rest, pMED, and pMS) sampled in each patient, 60-second filtered SEEG signals were divided into 12 non-overlapping 5-second epochs. The time series were then orthogonalized to minimize spurious correlations.^5^ Next, the AEC was calculated on each epoch and was averaged across them, thus obtaining an adjacency matrix of sFC for each subject and each of the three conditions. The size of the sFC matrices varied across patients, according to the number of analyzed SEEG leads (number of leads = 123.10 ± 26.82, mean ± standard deviation).

Connectivity matrices were represented through a graph, a mathematical object which allows to extract quantitative parameters to describe and summarize different topological properties of the network under study, such as centrality, integration, and segregation.^28^ A graph is defined by a set of nodes (in this case SEEG leads) and weighted edges connecting the nodes (*i.e.*, the value of AEC between each pair of leads). The static network topology during rest, pMED, and pMS was described using various graph theory indices, which characterize different topological network properties, as shown in Fig. 2. First, the global connectivity of the network was assessed by means of the global strength, which is the sum of all edges of the network that assess the extent of the interconnection between the different regions considered. Another evaluated network property was the centrality, which reflects the importance or influence of a node within the network. This property was measured through strength (the sum of all the edges of a node) and betweenness centrality (the extent to which a node lies on paths between other nodes). Moreover, the segregation, the degree to which a network can be divided into smaller, densely interconnected groups, was assessed using the clustering coefficient, which quantifies the tendency of a node’s neighbors to also be connected to each other. Finally, the integration, which indicates the network’s capacity to combine information across different regions, was evaluated using the characteristic path length. This metric represents the average shortest path between any two nodes and reflects how efficiently information is exchanged across the network. To avoid spurious effects due to differences in global strength among different networks, all metrics (apart from global strength) were calculated on graphs with normalized global strength.

**Figure 2.**
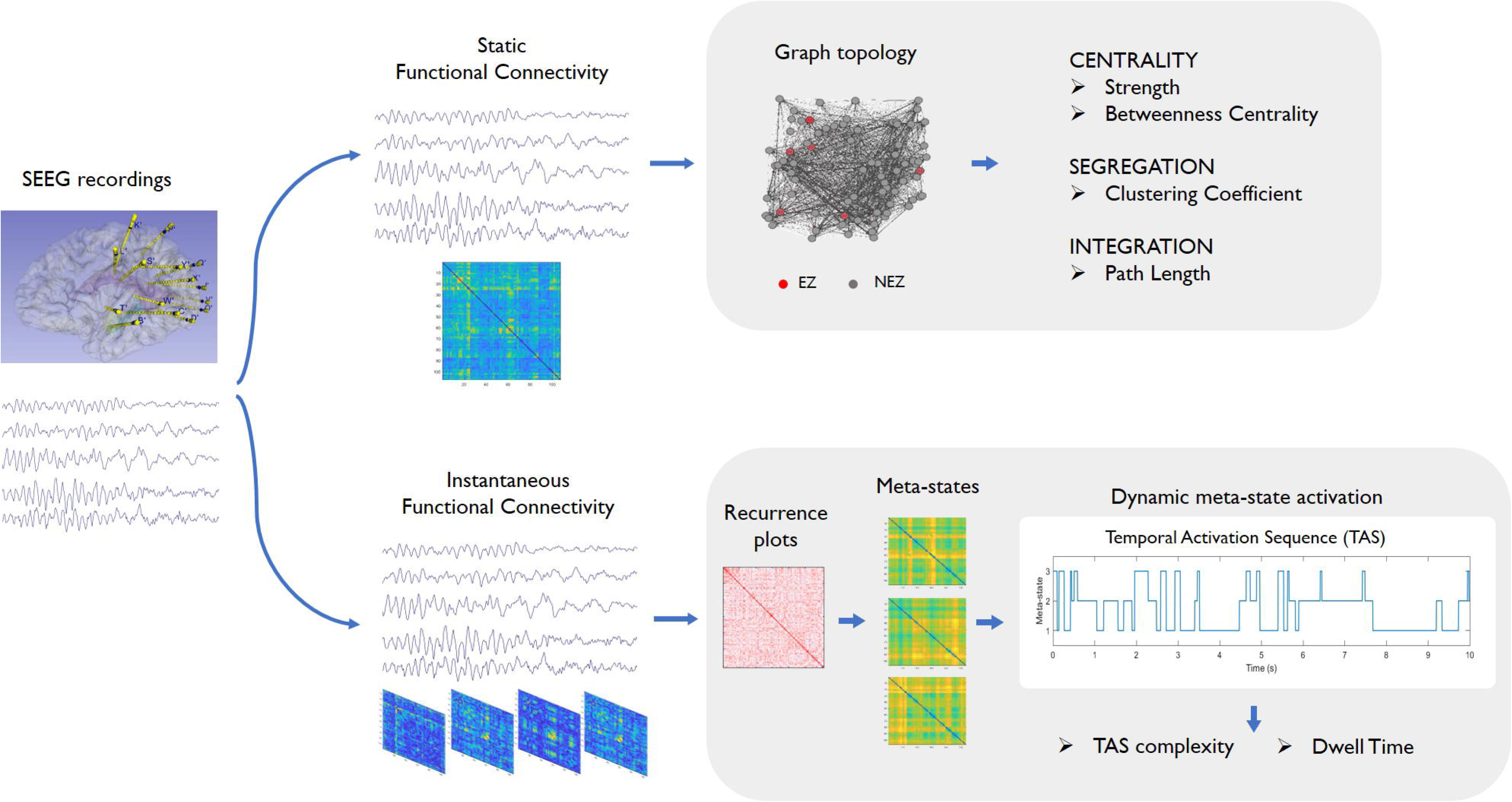
Methodological workflow. **Upper part** – static functional connectivity (sFC). The brain connectivity was estimated using a static approach by means of the orthogonalized AEC, and different network properties (centrality, segregation and integration) were assessed. **Lower part** – dynamic functional connectivity (dFC). The dFC was computed by: (i) calculating the instantaneous connectivity tensors by means of the IAC; (ii) computation of recurrence plots that assess recurrences in the connectivity matrices across time; (iii) identification of subject-based brain states recurrent in time, referred as “meta-states”, (iv) extraction of meta-state activation dynamics; and (v) estimation of dwell time y TAS complexity.

It is well known that the size of a network (*i.e*., the number of nodes and edges) can significantly influence its functional features.^29–31^ To address this issue, we applied a bootstrap approach following these steps:^29–33^ (i) Random downsampling of the graph’s nodes to the minimum common subset among patients, leading to a final network size of 48 nodes, 4 from the EZ and 44 from the non-epileptogenic zones (NEZ); (ii) *N_br_* = 100 bootstrap repetitions of the random downsampling; (iii) computation of each graph metric for each of the *N_br_* repetitions, followed by averaging across all repetitions; and (iv) averaging of the metric across all the leads within EZ and NEZ regions.

### 2.4. Dynamic Functional Connectivity (dFC)

The dynamic fluctuations of the network were assessed by means of a published method based on detecting recurrent network topologies over time, known as meta-states.^7^ These meta-states are considered as attractors of the neural network in each moment, governing their dynamic fluctuations across time. This methodology begins by computing the instantaneous amplitude correlation (IAC), a high temporal resolution version of the AEC (see Tewarie *et al*., 2019 for method details).^9^ For each of the three conditions (rest, pMED, and pMS), band-passed filtered SEEG signals were utilized to calculate the IAC between each pair of SEEG leads for each 60-s time sample. For each patient, we built a dynamic connectivity matrix of: [3 conditions x *N* leads x *N* leads x 15000 temporal samples (60 seconds · 250 Hz)], with the number of leads being different for each subject. Of note, before computing the IAC, the time series were orthogonalized to minimize volume conduction effects.^5^ After calculating instantaneous FC using IAC, we explored the temporal recurrences of connectivity matrices (*i.e.*, time slots in which the connectivity presents similar network topology) using recurrence plots (RPs) commonly used to visualize recurrent patterns in dynamical systems.^34^ RPs were 15000 x 15000 matrices, where the cell *C_ij_* (with *i,j* = 1,…,15000) indicates the topological similarity between the FC networks present in time samples *i* and *j*, calculated as the Spearman correlation between them. To reduce the computational cost of the methodology, we applied a data-driven windowing method^9^ that detects time periods where networks can be considered as stable. The IAC matrices were averaged in the time slots indicated by the above data-driven windows, and new (temporarily-constrained) RPs were constructed. This approach also presents the advantage of minimizing the effects of noise, as described in detail by Tewarie and colleagues.^9^

The RPs were considered as graphs, with each adaptive window being a node, and the correlation between them, the edges. Next, we applied the Louvain GJA community detection algorithm to the temporarily-constrained RPs to identify network topologies recurrent over time (communities),^35^ without *a-priori* defining the number of communities to detect. The Louvain GJA algorithm performs community segmentation maximizing modularity^36,37^ and reveals groups of time windows that are similar to others,^7^ The community detection was individually performed for each patient, as the different number and spatial locations of SEEG electrodes prevented the detection of group-representative states across subjects. To do this, we concatenated the windowed connectivity for the three conditions (rest, pMED, and pMS) to generate the RPs on which the Louvain GJA algorithm was applied. This allowed us to obtain the meta-states particular to each subject and their corresponding FC matrices.

Once the meta-states were obtained, we went back to the original, non-temporally aggregated connectivity matrices. Each temporal sample was assigned to a dominant meta-state by performing Spearman correlation of the FC matrix in that time sample with the FC topology of all the identified meta-states (*i.e.*, the meta-state displaying higher correlation with the network in that time sample was determined to be dominant for that time point). The symbolic time series indicating the dominant meta-state (*i.e.*, the assigned meta-state) in each sample during the 60-second epoch is called Temporal Activation Sequence (TAS).^7^ Derived from this metric, we employed two additional parameters:

**Dwell time** – This metric evaluates the average time the brain remains in the same dominant meta-state (Núñez et al., 2021). It is a widely used parameter in studies of dynamic brain state transitions, also known in the literature as “lifetime”.^38^

**TAS complexity** – To effectively capture the structural richness of the meta-state sequencing, we utilized Lempel-Ziv complexity (LZC).^7^ LZC relates to the number of distinct substrings and their rate of occurrence, with higher values indicating greater complexity.^39^ The LZC algorithm follows the method described by Abásolo *et al*. (2006),^39^ with the only difference being that the TAS is already a finite symbol sequence, so no conversion is required.

Finally, the whole process was repeated after removing the electrodes corresponding to the EZ, obtaining a NEZ network; and meta-states for each subject without EZ electrodes, as well as their corresponding TAS and ICT, were computed. Due to the reduced number of leads in EZ in some subjects, and the high complexity of these analyses, it was not possible to perform them exclusively on the EZ. The methodological procedure, including static and dynamic FC analyses, is illustrated in Fig. 2.

#### 2.4.1. Surrogate data for metric normalization

To determine whether the extracted measures represented genuine dFC rather than random fluctuations, we conducted surrogate data testing.^7,40^ To this end, we created surrogate versions of each EEG recording using the amplitude-adjusted Fourier transform (AAFT), a method that retains the amplitude distribution of the original time series.^41,42^ Importantly, we kept the same sequence of random numbers (uniform phase randomization) in all SEEG channels to maintain linear correlations and preserve static functional connectivity (FC).^40,42^

Following previous studies,^7,8,43^ all measures were normalized by dividing them by the average values calculated from 100 surrogate time series. This approach ensures that values close to one indicate behavior that can be attributed to random fluctuations, while values significantly different from one (either higher or lower) reflect behavior inherently associated with dFC.

### 2.5. Statistical analysis

The same statistical procedure was applied for both static (strength for the global network; and strength, betweenness centrality, clustering coefficient, and characteristic path length for EZ and NEZ regions) and dynamic metrics (TAS complexity and dwell time, for the global and the NEZ network). To avoid the assumption of normality and homoscedasticity of data, the non-parametric Friedman test was first applied to detect statistical differences among rest, pMED, and pMS conditions, for all frequency bands. To control type I error, false discovery rate (FDR) correction was applied to control for the number of bands and metrics.^44^

When a significant interaction was present, a post-hoc Wilcoxon signed rank test was performed to assess pairwise between-condition differences. A significance level of 0.05 was considered. Again, an FDR correction was used, so all *p*-values indicated in the results refer to the corrected ones. Signal processing and statistical analyses were performed using MATLAB (version R2018a and R2022a Mathworks, Natick, MA), with customs functions.

## 3. Results

### 3.1. Clinical findings

We considered 257 patients that underwent SEEG over a period of 17 years; of these, 39 (17 females – 44%) displayed both MED and MS preceded by at least 60 seconds of SEEG recording without any other pathological discharges. These 39 subjects were included in the study. We analyzed one MED and one MS for each patient. MEDs have a mean duration of 7,3 ± 13,3 (median: 3 seconds); MS have a mean duration of 56,3 ± 74,5 (median: 25 seconds). Fragmentary rhythmic interictal spike bursts lacking frequency or spatial evolution were not considered to be MED.

The mean age was 24.5 ± 12.0 years (mean ± standard deviation) at surgical treatment, and epilepsy duration of 14.6 ± 9.9 years (mean ± standard deviation). Structural lesions neuroradiologically identified and/or demonstrated after neuropathological evaluation were heterogeneous: seven focal cortical dysplasia (FCD) type I, five FCD type IIa, two FCD type IIb, six with no lesion at the MRI/histopathological examination, nine gliosis or scar, four periventricular nodular heterotopias, two cavernomas, two hippocampal sclerosis, one polymicrogyria with periventricular nodular heterotopia and one with oligodendroglial hyperplasia (MOGHE).

Postsurgical follow-up was 76.7 ± 41.8 months (mean ± standard deviation), range 17-174 months. Postsurgical seizure outcome at the last control was Engel class I in 30 patients (76.9 %) and Engel class II-IV in nine (23.1 %). All class II-IV patients had incomplete EZ resection for clinical reasons (functional area included in the EZ). Clinical information is summarized in Table 1.

**Table 1.**
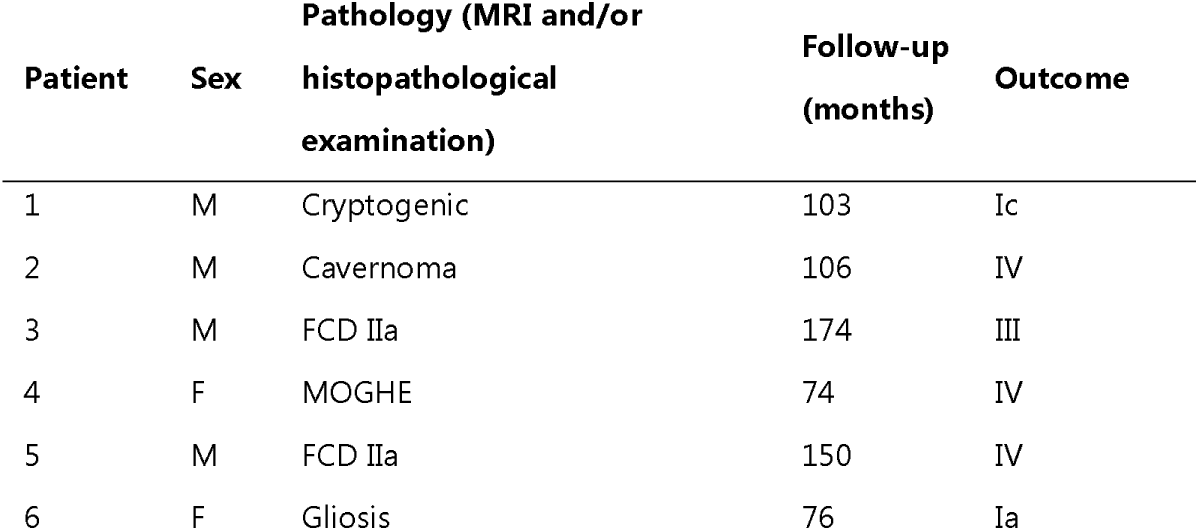
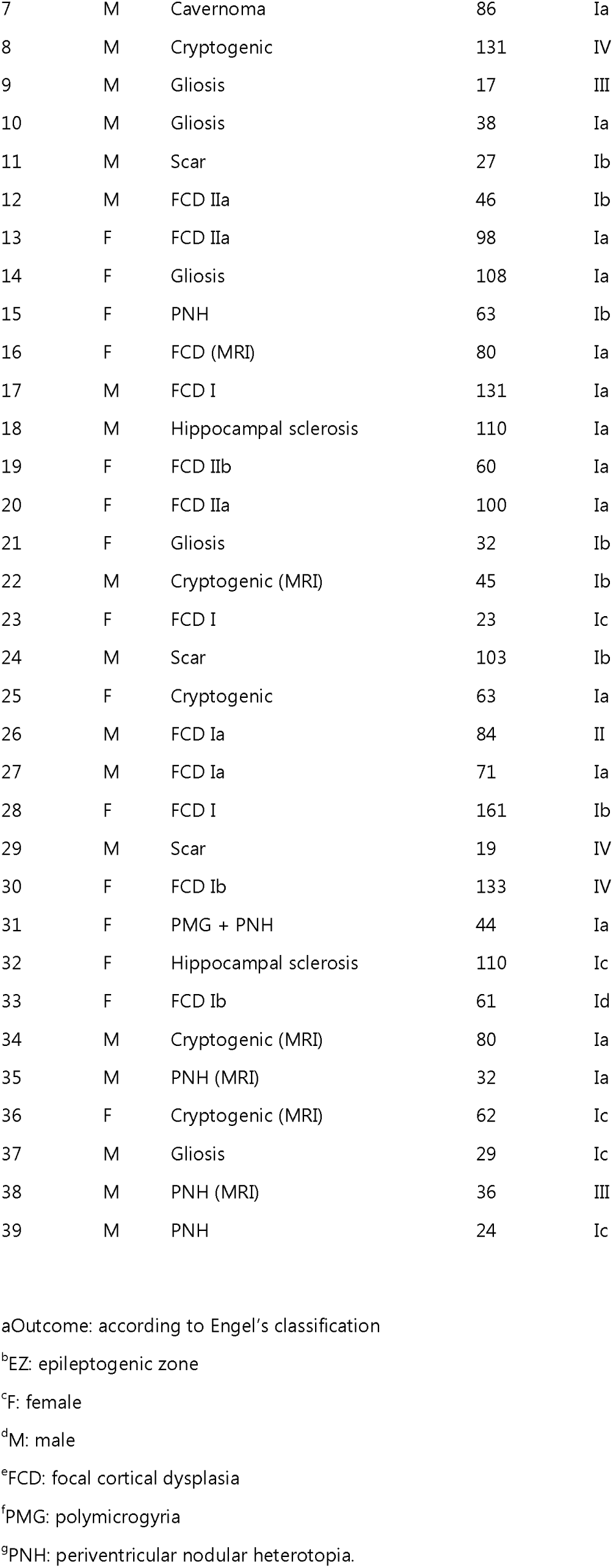
Clinical findings.

The EZ in our patient cohort could involve one or more than one lobe, with a prevalence of the frontal (*n* = 27) and temporal (*n* = 17) lobes, followed by the parietal (*n* = 14), occipital (*n* = 7), and insular (*n* = 6) lobes.

### 3.2. Static functional connectivity (sFC)

#### Network global strength

Fig. 3 shows the distribution plots of the global strength in different SEEG frequency bands in the three different conditions (rest, pMED, and pMS). Statistical results for Friedman test and post-hoc Wilcoxon signed-rank test are reported in Table 2. We found an interaction effect among the three conditions in all the frequency bands except for *theta*. For the same bands, post-hoc comparisons indicated an overall increase of global strength during pMED compared to rest. Moreover, pMED global strength was also significantly higher than pMS for the *beta* and *gamma* bands. No differences were found between rest and pMS.

**Figure 3.**
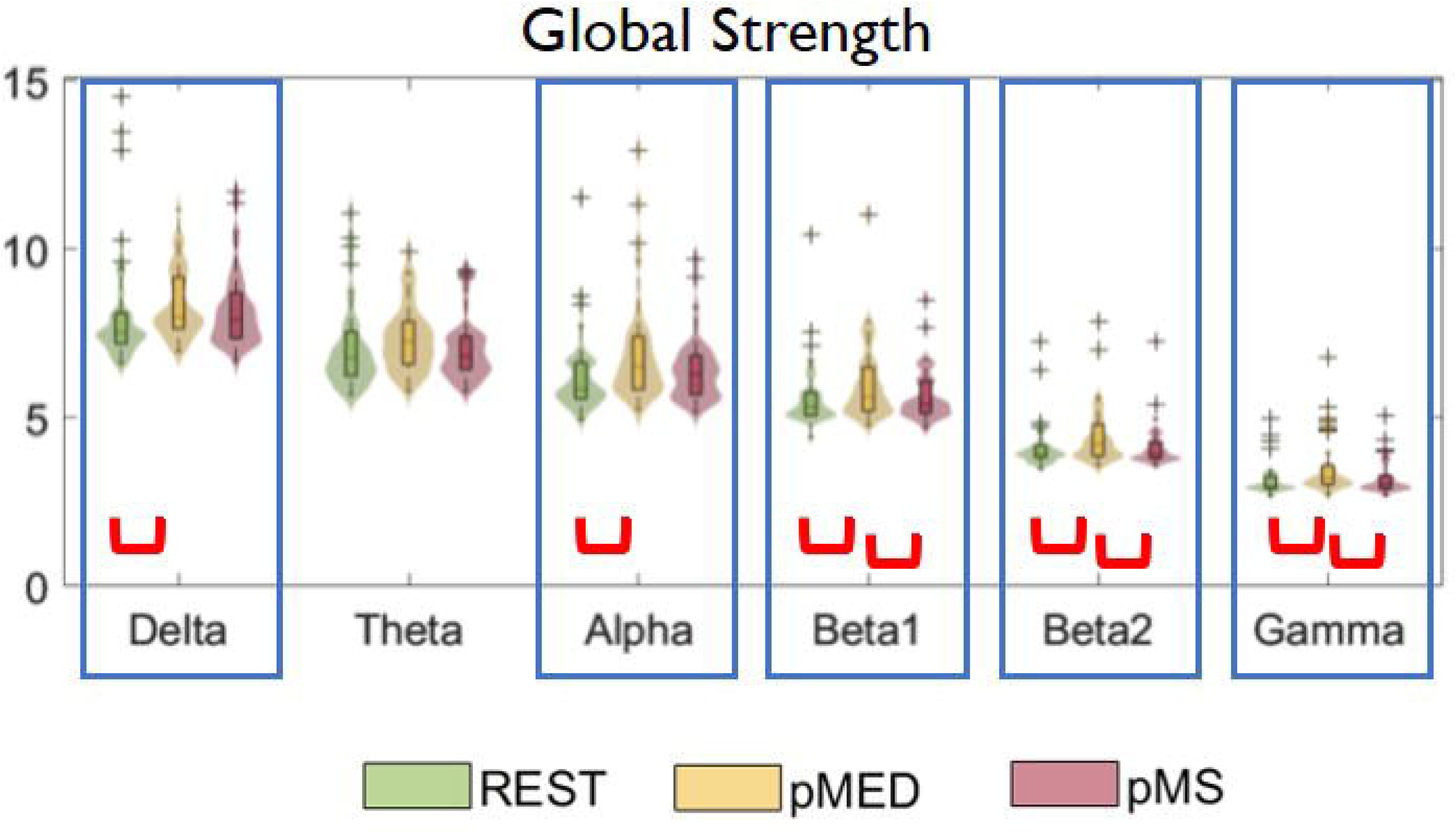
Distribution plots of the global strength estimated from static functional connectivity, for the three conditions (rest, preceding minor electrical discharges, pMED, and preceding major seizures, pMS). Between-group statistically significant differences are marked with blue rectangles (p < 0.05, Friedman test FDR-corrected). Post-hoc pairwise statistically significant differences are marked with red brackets (p < 0.05, Wilcoxon signed rank test FDR-corrected).

**Table 2.** Global strength.

| | | $\delta$ | $\theta$ | $\alpha$ | $\beta 1$ | B2 | $\gamma$ |
| --- | --- | --- | --- | --- | --- | --- | --- |
| **Friedman test** |  | 0,025 | 0,129 | 0,004 | 0,003 | 0,004 | 0,004 |
| **Post-hoc Wilcoxon signed rank** | rest-pMED | 0,044 | 0,092 | 0,012 | 0,017 | 0,000 | 0,000 |
|  | rest-pMS | 0,214 | 0,615 | 0,105 | 0,185 | 0,706 | 0,539 |
|  | pMED-pMS | 0,567 | 0,092 | 0,105 | 0,024 | 0,002 | 0,002 |

#### EZ and NEZ network topology

The distribution plots and the statistical comparisons among conditions for all the four remaining sFC metrics are summarized in Fig. 4, for both the EZ (left panel) and NEZ (right panel). Statistical values for the Friedman test and post-hoc Wilcoxon signed-rank test are reported in Table 3. In terms of nodal strength, Friedman test of EZ and NEZ showed similar results, with significant differences among the three conditions for all the frequency bands, except the NEZ *theta* band, with the most significant effect in the *beta2* and *gamma* for EZ. In the EZ, this difference was due to an increased nodal strength in pMED with respect to the rest in the *alpha* band, and increased values in pMED *beta1*, *beta2*, and *gamma* bands with respect to both rest and pMS conditions. The NEZ strength had a similar pattern, with the only exception of a significant lack of group interaction in the *theta* band.

**Figure 4.**
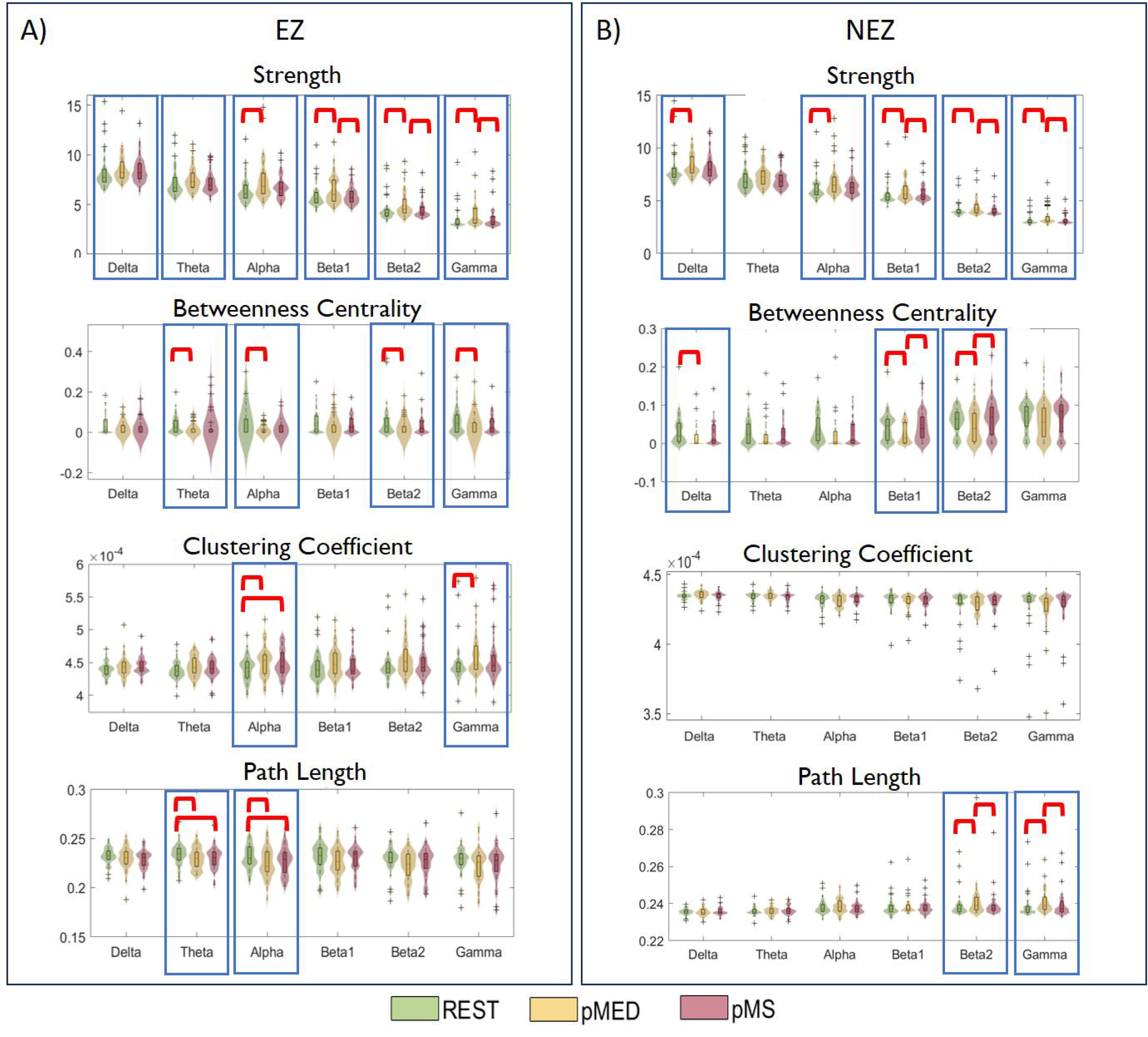
Distribution plots of the local static network metrics (strength, betweenness centrality, clustering coefficient, and path length) for both the epileptogenic zone, EZ (A) and the non-epileptogenic zone, NEZ (B) regions, and for all the frequency bands under study. Between-group statistically significant differences are marked with blue rectangles (p < 0.05, Friedman test FDR-corrected). Post-hoc pairwise statistically significant differences are marked with red brackets (p < 0.05, Wilcoxon signed rank test, FDR-corrected).

**Table 3.**
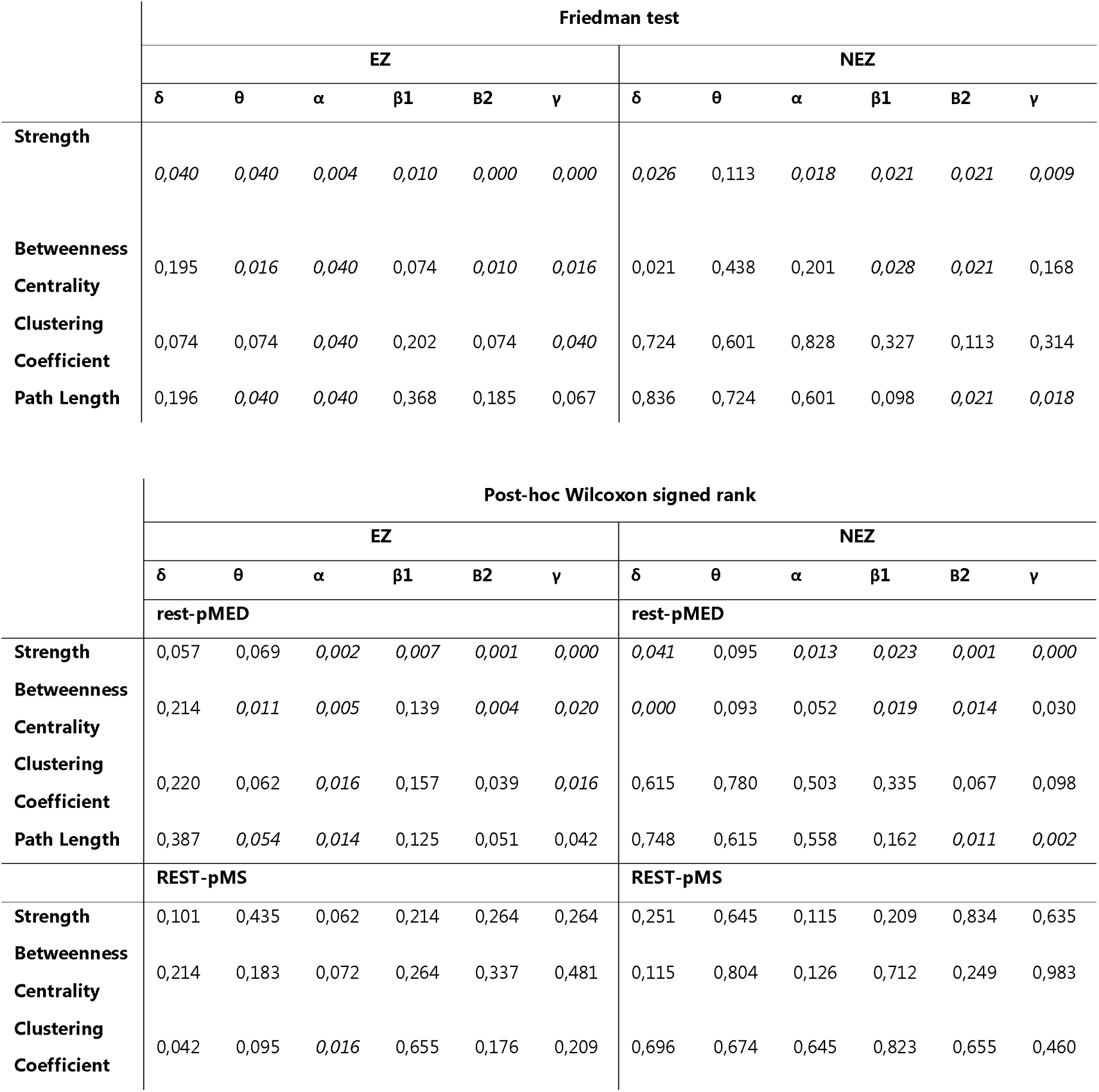
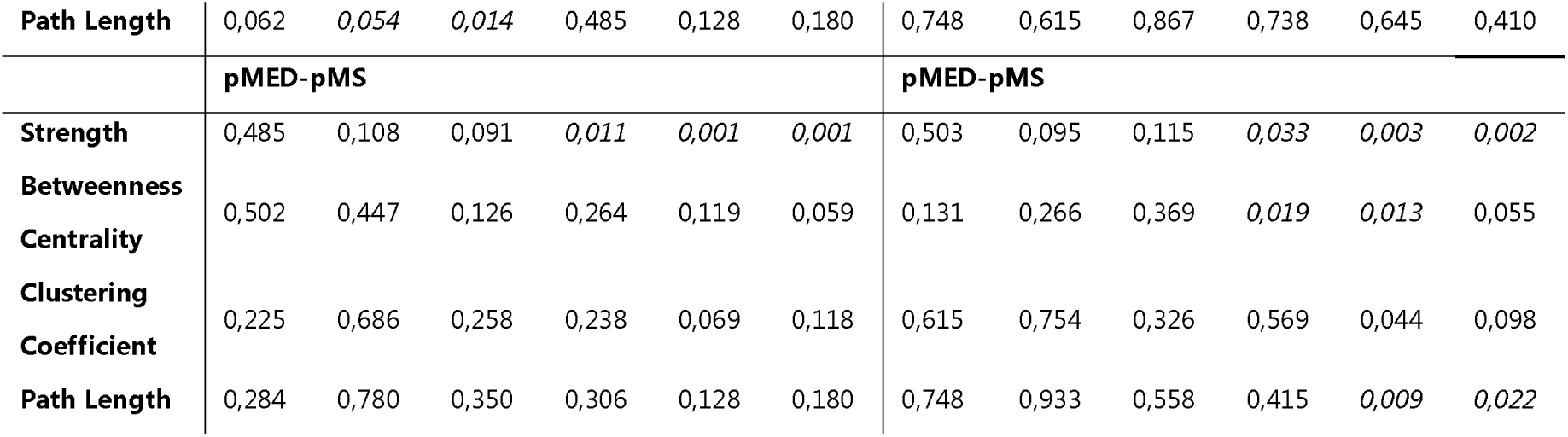
Strength, betweenness centrality, clustering coefficient, path length in the epileptogenic zone (EZ) versus non epileptogenic zone (NEZ).

Regarding the EZ, betweenness centrality alterations involved the *theta*, *alpha*, *beta2*, and *gamma* bands, with a significant reduction in pMED compared to rest. Furthermore, changes in EZ segregation for *alpha* and *gamma* bands were observed, due to higher clustering coefficient in pMED with respect to the rest condition in both bands and higher pMS *vs* rest values in *alpha* band. Lastly, regarding the path length, we identified statistically significant differences in *theta* and *alpha* bands, due to a reduced pMED and pMS path length compared to rest.

NEZ betweenness centrality alterations involved the *delta*, *beta1*, and *beta2* bands, with a significant reduced value in pMED with respect to rest and pMS. Contrarily to the EZ, no statistically significant differences between the three conditions in terms of clustering coefficient were found. Lastly, differences in path length were statistically significant in *beta2* and *gamma* bands, with higher path length during pMED than during both rest and pMS conditions. We found no differences in terms of clustering coefficients.

Overall, the results indicate the presence of network alterations differentiating pMED with respect to rest and pMS conditions, particularly in the *beta1*, *beta2*, and *gamma* bands, for all the topological properties (centrality, integration, and segregation). On the contrary, pMS showed a static network topology more like the rest condition, with only *theta* and *alpha* connectivity of the EZ indicating a network alteration during pMS with respect to the rest condition.

### 3.3. Dynamic functional connectivity (dFC)

To assess the dynamical patterns characterizing the three different conditions, two metrics of meta-state activation were calculated: TAS complexity and dwell time, both for the whole network (including EZ and NEZ regions) and for the NEZ network only. The meta-states for the EZ subnetwork could not be estimated due to the low number of EZ leads for each patient (ranging from 4 to 52), which made it impossible to obtain an accurate estimate of modularity. Fig. 5 shows the distribution plots and the statistical comparisons between conditions for TAS complexity (left panel) and dwell time (right panel), calculated on the whole network (Fig. 5a) and on the NEZ subnetwork (Fig. 5b). Statistical values for the Friedman test and *post-hoc* Wilcoxon signed-rank test are reported in Table 4. The overall results reveal an altered dFC in the *beta2* and *gamma* frequency bands for the pMED condition compared to both rest and pMS, primarily driven by NEZ subnetwork. This alteration is characterized by lower TAS complexity and higher dwell time in pMED relative to the other two conditions, indicating reduced dynamical variability in network reconfiguration during this state.

**Figure 5.**
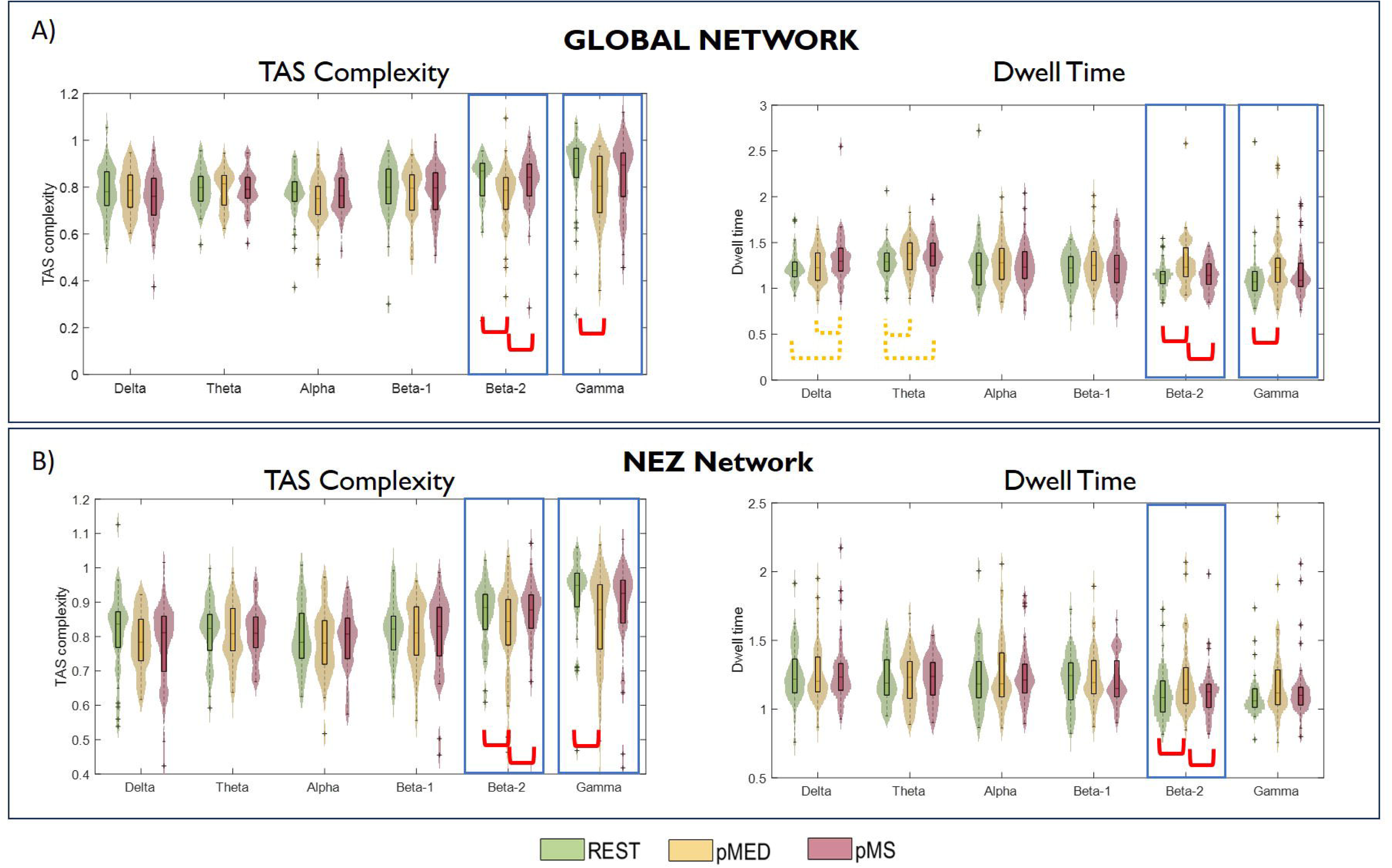
Distribution plots of the TAS complexity and dwell time for the global network (A) and the NEZ network (B). Between-group statistically significant differences are marked with blue rectangles (p < 0.05, Friedman test FDR-corrected). Post-hoc pairwise statistically significant differences are marked with red brackets (p < 0.05, Wilcoxon test FDR-corrected). Orange dotted brackets indicate a trend of difference that is not significant after FDR correction, even if close to significant effect (p < 0.06).

**Table 4.** (A) Dwell Time and (B) TAS Complexity in the whole network versus the non-epileptogenic zone (NEZ) subnetwork.

|  |  | **Dwell Time** |  |  |  |  |  |  |  |  |  |  |  |
| --- | --- | --- | --- | --- | --- | --- | --- | --- | --- | --- | --- | --- | --- |
|  |  | **Whole Network** |  |  |  |  |  | **NEZ subnetwork** |  |  |  |  |  |
|  |  | **$\delta$** | **$\theta$** | **$\alpha$** | **$\beta 1$** | **B2** | **$\gamma$** | **$\delta$** | **$\theta$** | **$\alpha$** | **$\beta 1$** | **B2** | **$\gamma$** |
| **Friedman test** |  | 0,175 | 0,116 | 0,174 | 0,199 | 0,008 | 0,035 | 0,926 | 0,882 | 0,882 | 0,664 | 0,022 | 0,174 |
| **Post-hoc** | rest-pMED | 0,780 | 0,067 | 0,308 | 0,316 | 0,008 | 0,007 | 0,738 | 0,748 | 0,615 | 0,592 | 0,022 | 0,026 |
| **Wilcoxon** | rest-pMS | 0,069 | 0,067 | 0,329 | 0,664 | 0,717 | 0,315 | 0,738 | 0,748 | 0,615 | 0,791 | 0,878 | 0,199 |
| **signed rank** | pMED-pMS | 0,069 | 0,834 | 0,738 | 0,738 | 0,008 | 0,314 | 0,738 | 0,748 | 0,615 | 0,592 | 0,022 | 0,145 |

|  |  | **TAS Complexity** |  |  |  |  |  |  |  |  |  |  |  |
| --- | --- | --- | --- | --- | --- | --- | --- | --- | --- | --- | --- | --- | --- |
|  |  | **Whole Network** |  |  |  |  |  | **NEZ subnetwork** |  |  |  |  |  |
|  |  | **$\delta$** | **$\theta$** | **$\alpha$** | **$\beta 1$** | **B2** | **$\gamma$** | **$\delta$** | **$\theta$** | **$\alpha$** | **$\beta 1$** | **B2** | **$\gamma$** |
| **Friedman test** |  | 0,700 | 0,498 | 0,251 | 0,794 | 0,004 | 0,046 | 0,836 | 0,836 | 0,285 | 0,552 | 0,034 | 0,005 |
| **Post-hoc** | rest-pMED | 0,823 | 0,856 | 0,125 | 0,748 | 0,008 | 0,006 | 0,501 | 0,878 | 0,664 | 0,845 | 0,053 | 0,008 |
| **Wilcoxon** | rest-pMS | 0,823 | 0,856 | 0,606 | 0,748 | 0,878 | 0,402 | 0,592 | 0,878 | 0,706 | 0,845 | 0,834 | 0,199 |
| **signed rank** | pMED-pMS | 0,583 | 0,856 | 0,321 | 0,748 | 0,017 | 0,172 | 0,856 | 0,878 | 0,664 | 0,845 | 0,026 | 0,092 |

#### Global Network (EZ+NEZ)

A significant interaction between conditions was found in the *beta2* and *gamma* bands for both TAS complexity and average dwell time. *Post-hoc* comparisons indicated statistically significant lower values of TAS complexity for pMED compared to rest. On the other hand, the post-hoc analysis of dwell time metric indicated an increased dwell time for pMED compared to rest in the same frequency bands. For both metrics, no significant differences between rest and pMS were found. However, pMS was characterized by higher dwell time than rest and pMED in the *delta* and *theta* frequency bands, with differences close to statistical significance (*p* = 0.06). Consistently with sFC results, lower frequency ranges appear to be associated with FC alterations leading to MS, while higher frequency being related to specific alterations characterizing pMED condition. This difference is not statistically significant after FDR correction, whereas showed a trend toward this direction.

#### NEZ Network

Considering only the NEZ subnetwork, the dFC results revealed the same pattern of differences than for the global network in the *beta2* and *gamma* band FC in pMED with respect to rest and pMS, with a reduced TAS complexity and higher dwell time in pMED. On the other hand, after removing the EZ, the trend of longer dwell time during pMS in the *delta* and *theta* range disappeared, suggesting a more specific role of the EZ low frequency functional connectivity in seizure dynamics. Of note, the statistically significant differences in dwell time in *gamma* also disappeared if the EZ was not considered.

## 4. Discussion

As reflected by the extensive literature in the field, epilepsy research has shifted from a local perspective toward a dynamic, complex, and whole-brain network standpoint, observing that the epileptic brain network experiences topological and functional alterations during interictal, preictal, and ictal stages.^45–47^ Understanding the dynamics of epileptogenic networks is of great clinical relevance, as it is essential for elucidating the mechanisms underlying seizure generation and propagation. In this study, we employed a comprehensive approach combining static and dynamic FC to investigate and differentiate the alterations in brain networks preceding MED and MS onset in patients with drug-resistant epilepsy. To our knowledge, this is the first study to quantify the difference between these two events within the same patient using SEEG recordings. We hypothesized that, according to the “pull-push theory”,^24,25^ this approach could offer a valuable model to better understand regulatory mechanisms of epileptogenic networks, and the interplay between synchronization and desynchronization mechanisms that can either produce or inhibit seizures. Our findings indicate distinct patterns of sFC and dFC preceding MED compared to both resting state and MS, suggesting the existence of potential inhibitory mechanisms involving the NEZ, which can suppress seizure occurrence and spreading in and from the EZ.

### 4.1. Static Functional Connectivity Analysis

Our analysis revealed significant alterations in network centrality, integration, and segregation properties, involving both the EZ and NEZ. Specifically, we identified an overall increased strength and a reduced betweenness centrality during pMED compared to resting state and pMS, particularly in the *beta* and *gamma* frequency bands, for both EZ and NEZ sub-networks. Strength and betweenness centrality are two complementary measures used to characterize the importance or centrality of nodes within a network. Strength quantifies the overall connectivity of a node in terms of number and strength of connections with other nodes, while betweenness centrality quantifies the extent to which a node acts as a bridge between different parts of the network.^28^ Their opposite trends (increased strength and reduced betweenness) suggest that the network’s nodes gain stronger local connections, while reducing their role as crucial bridges connecting different parts of the network. This scenario suggests a protective shift in the network structure, occurring only during pMED (and not during pMS), where nodes become more locally interconnected while reducing their role of critical hubs as connectors, thus being less sensitive to disruptions in specific regions. On the other side, the analysis of path length and clustering coefficient (measures of integration and segregation, respectively) revealed a different behavior between the EZ and NEZ sub-networks. The EZ shows a reduced path length and an increased clustering coefficient during both pMED and pMS compared to the rest condition. This reflects a more integrated network within the EZ, with higher modular organization and intragroup connections, while reducing the interactions with other regions. In contrast, the NEZ subnetwork exhibited a significant increase in path length during pMED, but not during pMS. This suggests that the NEZ sub-network, by reducing its integration with other brain regions, could play an inhibitory role in seizure propagation that controls the progression from MED to MS.

Altered integration and segregation properties have been extensively associated with epileptic networks. In a comprehensive review with meta-analysis, Slinger and colleagues^48^ reported that interictal structural networks of epileptic patients have a lower level of integration with respect to healthy controls. Previous studies also pointed out higher levels of segregation in epileptic patients, although this result is less homogeneous among studies.^48,49^ Our results highlight the distinct and opposite role of the EZ and NEZ in modulating integration and segregation prior to epileptic activity. Specifically, in the EZ, we observed increased integration and reduced segregation for both pMED and pMS, which may serve as mechanisms that predispose the network to seizure initiation. Conversely, the NEZ reduces network integration exclusively during pMED, but not during pMS, suggesting its role in suppressing the evolution of a MED into a MS.

Static connectivity alterations in epileptic networks have been widely studied,^13,50^ with inconsistent results possibly due to differences in methodological approaches as well as to the considerable localization or etiology variability in focal epilepsies, and the different criteria utilized to define the EZ. Most evidence suggests that resting state networks in focal DREs are associated with increased local connectivity in the EZ and decreased connectivity in widespread distant brain regions,^50^ with the EZ characterized by higher inward connectivity during resting state.^51,52^ These findings were confirmed by cluster partitioning analysis of intracerebral field responses evoked by intracerebral stimulation during SEEG,^53^ that demonstrated strong bidirectional connectivity between contacts included in the EZ. While epileptic networks have been largely associated with hyper-synchronization of brain activity, models of more complex interactions among hyper-synchronized and desynchronized sub-network nodes are being increasingly associated with seizure dynamics.^54–56^ Recent studies propose the existence of inhibitory networks mechanisms actively involve peri-EZ healthy brains regions that prevent the EZ from starting seizures. In a recent study, Johnson and colleagues^57^ postulated the interictal suppression hypothesis, a mechanism of seizure suppression in which the seizure onset zones are actively segregated by other healthy regions of the brain, suppressing their ability to connect with the rest of the network and initiate a seizure. The same authors reported a behavior of EZ during intracerebral stimulation, characterized by a relative decrease of *theta* band power coupled with a relative increase of *beta/gamma* band power when NEZs were stimulated.^57^ It has been long known that *theta* power is involved with long-range integration of brain regions, whereas *gamma* power is crucial for local integration.^58^ Thus, these findings suggest that EZ functional segregation is increased when NEZs are stimulated, underlying direct influence of NEZ on EZ.^57^ Accordingly, our results confirm statistically significant differences in EZ clustering coefficient in the *theta-alpha* range in the rest condition with respect to pMS and pMED, supporting the hypothesis of a higher EZ segregation before seizure onset.

Narasimhan *et al*., 2020^52^ highlighted high inward connectivity in the EZ reflecting inhibitory input from other regions to prevent seizure activity, which may flip the direction when seizure activity begins. Similarly, Gunnarsdottir and colleagues^59^ developed a ‘source-sink’ metric to accurately identify the EZ, based on the hypothesis that when a patient is not having a seizure, the EZ ‘sink’ nodes is inhibited by neighboring regions (the ‘source’ nodes). Our theoretical scenario shows an EZ similarly organized (and thus, similarly prone to generating seizures) in both conditions (pMEd and pMS), while NEZ is organized differently in pMED and pMS. Our findings appear coherent with the previous ‘inhibitory hypothesis’ but using a novel model to discuss the differential capacity of the brain to suppress or permit seizures before MS and MED (not only during the rest).

### 4.2. Dynamic Functional Connectivity Analysis

Dynamic functional connectivity analysis allowed us to explore the temporal evolution of brain FC meta-state organization preceding MED and MS onset. Meta-states analyses revealed distinct temporal patterns of network activation during different conditions, with lower TAS complexity and longer dwell time observed during pMED compared to both rest and pMS conditions, particularly in high frequencies activity bands. The reduction in TAS complexity suggests a more constrained and less variable network state preceding MED onset. Furthermore, longer dwell times during pMED indicate longer period of persistence in meta-states, reflecting prolonged periods of network stability. This pattern did not vary when considering either the whole network or the sub-network composed only by the NEZ. Overall, our results suggest inhibitory mechanisms activated before MED, based on the reduction of the intrinsic dynamic network fluctuation that characterizes epileptic networks. Interestingly, meta-state dFC analysis did not reveal a different pattern preceding seizures (*i.e.*, in pMS), except for a trend characterizing the EZ role in low-frequency ranges to increase the stability of meta-states (longer dwell time) before seizure onset. This could suggest an opposite mechanism facilitating seizures. However, the method applied, based on similarity recurrence plots, does not allow to properly study the specific EZ dynamics, due to the low number of nodes composing this sub-network preventing a reliable estimation of network similarity across time.

Dynamic FC has been recently developed and employed to characterize the alterations that schizophrenia, mild cognitive impairment, and Alzheimer’s disease elicit in meta-states topology and their dynamical transitions in auditory oddball task and resting state scalp EEG recordings.^7,8^ The study of dynamic epileptic networks is relatively recent and most of the studies in literature are based on fMRI data, which cannot provide information about fast temporal neural oscillations. In this framework, some recent studies suggested that epileptic patients have higher fluctuation of resting state FC over time compared to healthy controls,^60,61^ thus suggesting that temporal stability/instability of network organization is a useful biomarker of ictogenesis. Khambhati *et al*., (2015)^62^ defined different brain statuses, associated with different connectivity configuration during pre-ictal, ictal, and post-ictal stages. Their results showed that epileptic networks are characterized by hubs of persistent strong connections surrounded by rapidly reconfiguring weak connections that, preceding seizure generation, benefit from high network flexibility to drive seizure generation through a rapid reorganization of weaker connections in the epileptic network.^62^ In a further study, the same team proposed a “push-pull control” mechanism on virtually resected epileptic brains.^24^ According to this model, desynchronizing and synchronizing nodes act as antagonists’ determinants of the spread – or confinement – of dynamic processes. Starting from electrocorticography data, the authors demonstrated that synchronizability in the high-*gamma* networks is increased before seizures that spread compared to seizures that remain focal. Also, with an innovative method of virtual cortical resection, they identified synchronizing and desynchronizing brain regions. These control regions act differently in the pre-seizure and seizures epochs, especially in the area surrounding the EZ. In line with this evidence, our overall results suggest a protective mechanism based on the dynamic reconfiguration of the network, especially the NEZ, into a less flexible and less dynamically variable network able to influence (or even prevent) seizure occurrence.

### 4.3. Limitations and future research lines

This study has several limitations that warrant discussion. Regarding the meta-state analysis, a deeper understanding of the specific impact of the EZ on state dynamics would be valuable. However, in some subjects, the limited number of SEEG electrodes prevented accurate estimation of functional connectivity (FC) network self-similarity when focusing solely on EZ electrodes, thereby hindering the application of the Louvain algorithm for community detection. Addressing this challenge could involve methodological advancements, such as performing source estimation on the SEEG data to create a full cortical source map of SEEG activations.

Additionally, while the average dwell times and TAS complexity provide a broad overview of meta-state dynamics, future research could delve into the specific timing of state activations. Such an approach could facilitate a more detailed per-subject analysis of state dynamics and their potential connection to inhibitory mechanisms that might prevent seizures in the pMED condition compared to pMS. Another limitation is that this study focused only on the period preceding MED and MS, without analyzing seizure progression. This constraint arose because the dFC method employed requires consistent time-window comparisons across events of interest, and MED and MS differ in duration. Future research could explore alternative approaches to analyze seizure progression dynamics despite these differences. Furthermore, while temporal resolution is excellent, SEEG shows limited spatial resolution and implantation is patient-dependent. It must also be considered that our evaluation is aimed at patients with focal seizures, nothing can currently be inferred about generalized epilepsies which represent another interesting population to be evaluated in future studies. Finally, the relationship between sFC and dFC results and the underlying pathology was not analyzed, nor was the effect of antiseizure medications used by the patients during recordings.

## 5. Conclusions

This study explored the differences in brain networks preceding pMED and pMS in patients with drug-resistant epilepsy, using a comprehensive approach that combines static and dynamic functional connectivity. Our findings highlight distinct patterns of brain activity that precede pMED compared to resting state and pMS, suggesting the existence of inhibitory mechanisms involving NEZ capable of suppressing the onset and spread of seizures from EZ.

Overall, our results suggest that the brain network structure dynamically reconfigures to either hinder or facilitate seizures. The NEZ, in particular, appears to play a crucial protective role by stabilizing the network and preventing the spread of seizures. These insights pave the way to better understanding and treatment of epilepsy by targeting these dynamic network changes.

## Abbreviations

AEC: amplitude envelope correlation
dFC: dynamic functional connectivity
DRE: drug-resistant epilepsy
EZ: epileptogenic zone
FC: functional connectivity
FCD: focal cortical dysplasia
fMRI: functional magnetic resonance imaging
IAC: instantaneous amplitude correlation
ICT: instantaneous correlation tensor
MED: minor electrical discharge
MEG: magnetoencephalography
MS: major seizure
NEZ: non-epileptogenic zone
pMED: pre-minor electrical discharge
pMS: pre-major seizure
RP: recurrence plot
SEEG: stereo-electroencephalography
sFC: static functional connectivity
TAS: temporal activation sequence

## Acknowledgments and funding

This research was funded: (i) by EPICARE project of Associazione Paolo Zorzi per le Neuroscienze, Italian Ministry of health, Grant/Award Number: RF-2016-02362195; (ii) by grant PID2022-138286NB-I00 funded by “Ministerio de Ciencia e Innovación/Agencia Estatal de Investigación/10.13039/501100011033/”, by “ERDF A way of making Europe”; (iii) and by “CIBER en Bioingeniería, Biomateriales y Nanomedicina (CIBER-BBN), Spain” through “Instituto de Salud Carlos III” co-funded with ERDF funds. PN was supported by the ERA-Net FLAG-ERA JTC2021 project ModelDXConsciousness (Human Brain Project Partnering Project).

## Supplementary material

No supplementary materials are available for this manuscript.

## Data availability

The data that support the findings of this study and the codes for the analysis are available from the corresponding author upon reasonable request.

